# Identification of Cancer Risk Groups through Multi-Omics Integration using Autoencoder and Tensor Analysis

**DOI:** 10.1101/2023.09.12.23295458

**Authors:** Ali Braytee, Sam He, Shuxian Tang, Yuxuan Sun, Xiaoying Jiang, Xuanding Yu, Inder Khatri, Mukesh Prasad, Ali Anaissi

## Abstract

Identifying cancer risk groups by integrative multi-omics has attracted researchers in their quest to find biomarkers from diverse risk-related omics. Stratifying the patients into cancer risk groups using genomics is essential for clinicians for pre-prevention treatment to improve the survival time for patients and identify the appropriate therapy strategies. This study proposes an integrative multi-omics framework that can extract the features from various omics simultaneously. The framework employs autoencoders to learn the non-linear representation of the data and applies tensor analysis for feature learning. Further, the clustering method is used to stratify the patients into multiple cancer risk groups. Several omics were included in the experiments, namely methylation, somatic copy-number variation (SCNV), micro RNA (miRNA) and RNA sequencing (RNAseq) from two cancer types, including Glioma and Breast Invasive Carcinoma from the TCGA dataset. The results of this study are promising, as evidenced by the survival analysis and classification models, which outperformed the state-of-art. The patients can be significantly (p-value<0.05) divided into risk groups using extracted latent variables from the fused multi-omics data. The pipeline is open source to help researchers and clinicians identify the patients’ risk groups using genomics.

Additional Key Words and Phrases: Multi-omics, Autoencoders, Tensors, Cancer risk groups

## 1 INTRODUCTION

The subdivision of cancer and the identification of risk groups are of great significance in medicine for the diagnosis and treatment of cancer. Currently, in clinical practice, cancers are commonly treated according to their histological origin and pathological features. This approach has some limitations, such as similar histopathological features in some tumour masses, but their clinical presentation is quite different and corresponds to different risk groups. Several studies [Lee and Kim(2021)] have shown that the pathological system of tumours at the molecular level is well characterised in terms of their parthenogenesis and stage of development. Fortunately, as the Human Genome Project progresses and new sequencing technologies continue to emerge and spread, a wealth of omics data is being generated that contributes to a better understanding of the issues involved. Nevertheless, due to the inherent complexity of biological systems, there is a limit to the information provided by a single piece of omics data. Genomic variation caused by somatic mutations, epigenetic changes, individual differences and environmental influences are possible during tumour development. The traditional analyses based on individual omics cannot capture the heterogeneity of all biological processes [Chaudhary et al.(2018)]. On the other hand, using omics data also poses statistical modelling and computational challenges. In some omics data, there is the problem of a small number of samples and a large number of features [Braytee et al.(2017)]. Further, reliance on a single-omics may lose much information due to high data dimensionality and low sample size challenges. Therefore, these problems with single-omics hinder the better identification of risk groups or clinical phenotypes.

Recently, there has been a growing trend towards studying and analyzing multi-omics data, including genomics, epigenomics, transcriptomics, proteomics, metabolomics, microbiomics, imaging, and others. The use of integrated data analysis has various advantages. It compensates for the lack of information in single-omics data and provides an integrated view of cancer analysis at the molecular level. This approach can play an essential role in assessing metastasis and selecting treatments for patients, thus contributing to the development of precision medicine. Few studies related have used autoencoders in deep learning to extract features of multi-omics data and use these new features to build predictive models [Chaudhary et al.(2018)] [Ding et al.(2018)]. Further, some studies have used unsupervised feature extraction for multi-omics based on tensor decomposition [Taguchi(2017)] [Taguchi(2019)]. However, the small size omics datasets have not been considered and identifying the risk groups from multiple omics data has not been investigated.

This study develops a multi-omics feature learning framework to stratify patients into high and low-risk groups by minimising information loss and learning significant features. Autoencoders are used as a dimensionality reduction method to capture the non-linear relationships between the data to maximise the retention of the original information in each single-omics data. Then, the latent variables of each omic are concatenated, and further feature learning is carried out using tensor analysis. Combining deep learning and tensor analysis avoids overweighting omics datasets due to high dimensionality while learning important common features across multi-omics. Our proposed framework comprises three main components. Firstly, the original omics data is dimensionalised using autoencoders, which employ a combination of non-linear functions to reconstruct the original input. It is known that this method performs well when applied to biological data, with less information lost [Chaudhary et al.(2018)] [Ding et al.(2018)] [Zhang et al.(2018)] [Yao et al.(2022)] [Zhou et al.(2022)] and is therefore well suited to handle omics information. Secondly, the processed multi-omics data is fused into a tensor. The significant global features of different omics datasets can be learned through CANDECOMP/PARAFAC (CP) decomposition to extract interpretable latent factors. While the original data may not be fully recoverable from the CP decomposition of the compressed data, we focused on obtaining a more interpretable and meaningful representation of the data that captures its essential characteristics. Finally, the components extracted from tensor decomposition are utilized for clustering. The clustering results are evaluated using survival analysis. Additionally, a supervised learning model is built and used to predict Tumor Purity for the breast dataset due to the availability of class labels.

The practical relevance of the results generated by the proposed framework is evident. Specific risk groups could be detected earlier based on the framework results, which help clinicians to choose more appropriate therapies at different stages of treatment. Meanwhile, tensor analysis of multi-omics combined with deep learning methods may inspire more ways to identify cancer risk groups from the molecular level. Our contributions are summarized as follows:

- We propose a non-linear multi-omics method that considers the non-linear relationships between features in the assays.
- We integrate Tensors in the proposed model to extract expressive feature sets that capture important patterns and relationships in the data.
- We thoroughly evaluate our methods on two public datasets: Glioma and Breast Invasive Carcinoma. Our results are highly promising, as the survival analysis and classification models indicate.

## 2 RELATED WORK

With the development of high-throughput sequencing technology, omics analysis techniques are becoming increasingly mature and sophisticated. In contrast, the integration and analysis of multi-omics data have become a new direction for researchers to explore the mechanisms of life. At the same time, various challenges exist in the fusion analysis of multi-omics, such as high dimensional data with a relatively much smaller sample size and heterogeneity of individual omics. This work focuses on the tensor analysis of multi-omics data. This section outlines the main research findings in multi-omics, and the relevant methods involved in the processing and analysis of multi-omics data are reviewed.

### 2.1 Multi-omics Analysis

Many studies were conducted early to analyse a single-omics data [Conesa et al.(2016)]. However, the complexity of biological systems cannot be fully characterized by a single-omics. For example, while genomics has revealed genetic alterations in cancer patients, not all genetic variants cause changes in their expression and function [Long and Wang(2020)]. Furthermore, because of the high levels of noise that can be generated in omics data, experiments performed in isolation may lack the statistical significance to reveal valuable correlation results. Millions of single nucleotide variants (SNV) may be identified in a typical genome-wide association, and it is difficult to determine which SNV is the actual cause of a disease. Thus, studying biomolecular changes in only one aspect makes it difficult to understand complex biological processes, particularly salient in complex diseases.

Several studies have devoted considerable effort to investigating how to address the complex multi-omics data. Specifically, A study integrates complementary data from different perspectives, such as the genome, transcriptome, proteome, metabolome and interactome to reduce noise and boost statistical power [Ideker et al.(2011)]. Cohen et al. (2018) combined cell-free DNA mutations and circulating protein biomarkers to develop a new blood-based prediction method, CancerSEEK, which enables early diagnosis of cancers and locates the organ of origin of these cancers [Cohen et al.(2018)]. Tepeli et al.(2019) use somatic mutation, transcriptomics and proteomics data to find kidney cancer subgroups. The integrated analysis of multi-omics data has brought great help in understanding complex biological systems, offering great possibilities to understand the molecular regulatory mechanisms in biological systems, the mechanisms of gene expression regulation and even to simulate the natural conditions of biological systems [Tepeli et al.(2020)]. Yet few studies use multi-omics to predict risk groups for specific cancers.

### 2.2 Integration Approaches of Multi-omics data

Data integration in multi-omics combines data from different technologies or sources to provide the maximum amount of valuable information. Depending on the timing of integration, data integration can be broadly classified into three types: early, intermediate and late. The basic idea of early integration is to join the different data from the original or dimension reduction process into a single large matrix. Then, it is fed into a model to obtain prediction results [Zitnik et al.(2019)]. It can consider correlations between features as long as the data are not redundant. However, it ignores the unique distribution of each histological data type, the weights need to be normalised, and the dimensionality of the input data is increased. Therefore, it is essential to find ways to mitigate these issues when integrating multi-omics data using early integration methods.

Intermediate integration is the joint integration of multi-omics by preserving the data structure of the dataset without prior transformation and without relying on simple joins. It is an algorithm that fuses them through a joint model. This method reduces the complexity of multi-omics datasets and can address the problem of dataset diversity [Rappoport and Shamir(2018)]. In the EL-Manzalawy et al. (2018) study, to reduce the loss of information caused by selecting features for each group individually, they used the mRMR extension as an intermediate feature selection method that selects features by considering complementarity within and across histological blocks [El-Manzalawy et al.(2018)]. This integration is highly performed, but the heterogeneity between datasets can prevent this integration from working correctly. Only a few methods can discover patterns shared between omics, requiring the development of more new algorithms to integrate data. Late integration involves training separate models for each omics data to learn features combined as input to a classifier or regressor [Sharifi-Noghabi et al.(2019)]. This approach avoids some of the challenges of collecting diverse data using specialized tools for each omics type. However, this strategy only integrates predictions for each omic independently, and the cost of extracting features to integrate is high. Additionally, it fails to capture omics interactions and prevents models from sharing knowledge or exploiting complementary information during the learning process. As a result, combining predictions needs to be precise to leverage multi-omics data and gain insight into the underlying biological mechanisms of disease, leading to lower reliability [Picard et al.(2021)].

### 2.3 Dimensionality Reduction for Multi-omics

A study integrates several types of statistical methods for gene-wide measures to predict skin melanoma prognosis with the help of dimensionality reduction techniques, including elastic net, sparse PCA (sPCA) and sparse PLS, which are used to extract variables from multi-omics data [Jiang et al.(2016)]. Park et al. (2020) first identified homogeneous blocks of variables and then used sPCA on each omics dataset to extract the sPCs. Multi-omics factor analysis (MOFA) can be considered a statistically rigorous generalisation of (sparse) PCA to multi-omics data. In the study by Argelaguet et al. (2018), MOFA enabled the inference of a set of (hidden) factors to capture the biological and technical sources of variability [Argelaguet et al.(2018)]. The acquired factors supported subsequent analyses such as identifying sample subgroups, data interpolation and abnormal samples. There are considerable constraints to MOFA and related factor models, involving their extendibility and lack of ability to interpret ancillary information about inter-cellular structure [Argelaguet et al.(2020)]. Recently introduced in bioinformatics, deep learning techniques have also been used for multi-omics data-related tasks, which can perform well with dimensionality reduction. Autoencoder (AE) is one of the data compression algorithms and is divided into two parts: the first part is the encoder, which is generally a multi-layer network that compresses the input data into a vector (also known as a bottleneck layer), thus reducing the dimensionality. The second part is the decoder, where the bottleneck layer is reconstructed into the same data as the original input after passing through a multi-layer network. Chaudhary (2018) extracted compacted features from liver cancer data using autoencoder to process RNA, miRNA, and DNA methylation data. A Cox model was used to filter the compacted features based on survival time to obtain samples with new features [Chaudhary et al.(2018)]. The samples were clustered to obtain labels, and the SVM classification model was trained using label supervision. Ding et al. (2018) used deep learning methods to identify informative features in the multi-omics data. The classifier trained on this basis could predict the effectiveness of drugs in cancer cell lines. They used autoencoder to effectively reduce the dimensionality of the integrated data [Ding et al.(2018)]. For multi-omics cancer datasets with high dimensionality and noise, these deep learning-based multi-omics integration methods have advantages over traditional methods due to the strong fitting ability of deep neural networks. Therefore, our study will use an autoencoder to reduce the dimensionality of the omics data. Unlike Chaudhary (2018) and Ding et al. (2018), we will not use an autoencoder for the fused data because of the concern that data with too much dimensionality will overwhelm smaller data, but instead use autoencoder for each technology separately for dimensionality reduction to ensure that the information of each omic is maximally retained. Several methods have utilized autoencoders for dimensionality reduction by concatenating multi-omics datasets into a single, two-dimensional matrix. However, unfolding the data and analyzing it using two-way methods can result in information loss, as it disregards the modular structure that is inherent in the data [Lemsara et al.(2020)], [Ma and Zhang(2019)], [Zhang et al.(2018)], [Song et al.(2022)], [Chaudhary et al.(2018)].

### 2.4 Tensor Decomposition

An unsupervised feature extraction method based on tensor decomposition is proposed by creating k pattern tensors based on a multi-view dataset, then applying HOSVD to decompose these tensors and finally performing a clustering algorithm [Taguchi(2019)]. By measuring gene expression in combination with RNA-seq analysis in adipose, lymphoblastoid cell lines (LCL) and skin, Hore et al. (2016) decompose 3D tensors of gene expression using the Bayesian sparse tensor decomposition model to reveal the gene networks associated with genetic variation [Hore et al.(2016)]. The CANDECOMP/PARAFAC (CP) decomposition is applied to genomic and epigenomic data. After decomposing the tensor, learning is achieved using Support Tensor Machine Regression (STR) and Ridge Tensor Regression (RTR). Using such an approach, the CP decomposition constraint performs more robust than models based on individual data types and concatenation methods [Fang(2019)]. Fanaee-T and Thoresen (2019) use integrated dimension reduction and tensor decomposition in their implementation of visualising multi-omics data [Fanaee-T and Thoresen(2019)]. In addition, Taguchi and Turki (2021) propose kernel tensor decomposition (KTD) to improve multi-omics analysis [Taguchi and Turki(2022)]. Jung et al. (2021) propose a Multi-Omics Non-negative Tensor decomposition for Integrative analysis (MONTI), which uses a three-dimensional tensor with the addition of non-negative constraints when exploring subtype-specific features such as breast cancer and other clinical features for decomposition [Jung et al.(2021)].

In conclusion, the literature suggests that several methods have been proposed for analyzing multi-omics data. However, there are still several open challenges that need to be addressed. These include addressing the variation in the dimensionality of different omics data, accurately representing non-linear data, identifying cancer risk groups using information from various omics, and retaining the maximum amount of information from omics features during dimensionality reduction before data integration.

## 3 METHODS AND MATERIALS

### 3.1 Data Collection

The data used in this study were collected from an open platform LinkedOmics [Vasaikar et al.(2018)], which provides access to multi-omics data from all 32 TCGA Cancer Types and 10 Clinical Proteomics Tumor Analysis Consortium (CPTAC) cancer cohorts. To ensure enough samples to support our training and testing, we selected the Glioma and Breast Invasive Carcinoma cancer types as they have more than 1000 samples available. Four omics data have enough samples and significant differences in the size of the features for each cancer type. The four omics selected for breast are methylation (CpG-site level, HM450K), miRNA (HiSeq, Gene level), RNAseq (HiSeq, Gene level), and SCNV (Focal level, log-ratio). Similarly, in glioma, the same omics are chosen except for changing the miRNA to miRNA (Gene level) as there is no miRNA (HiSeq, Gene Level) in the available data. All the omics contain continuous data only as researching on mixture data type is out of scope for this study. The selected omics data include portions of the shared samples across the four technologies and clinical information. To ensure that the same set of common samples was used in the experiments, we matched each of the four technologies’ data with the corresponding clinical data to define our dataset. Consequently, we obtained 616 common raw samples for breast cancer and 508 for glioma..

The dimensions of the omics data varied significantly, with the largest being methylation, which had up to 335,854 dimensions, and the smallest being SCNV with only 69 dimensions. RNAseq had 20,155 dimensions and miRNA had 823 dimensions. The huge dimension difference makes handling data loss and delusion during dimension matching challenging. All the values in omics are continuous data. Further, we noticed many features only contained values for a few samples while all others were zero. The collected omics data of both cancer types have two common challenges. The first challenge is the huge size difference between different omics. For example, SCNV has only 69 genes, while methylation can have more than 330 thousand genes. This difference does not allow combining the data because the larger ones may dilute the lower-size omics. The other challenge is the low number of samples after the common samples are selected across various omics and the huge number of dimensions in some technologies, such as methylation.

### 3.2 Our Framework

Our proposed framework consists of three main components, as shown in Figure 1. In the first component, meaningful features are extracted for each omic using separate stacked autoencoders. In the second component, the output as extracted features of each omic is fused by a 3D tensor which stacks three same size matrices (*A, B, C*). The tensor is decomposed using CP decomposition. Finally, using decomposed factors from the tensor to generate the risk prediction by using different kinds of prediction models such as classification and clustering.

**Fig. 1.**
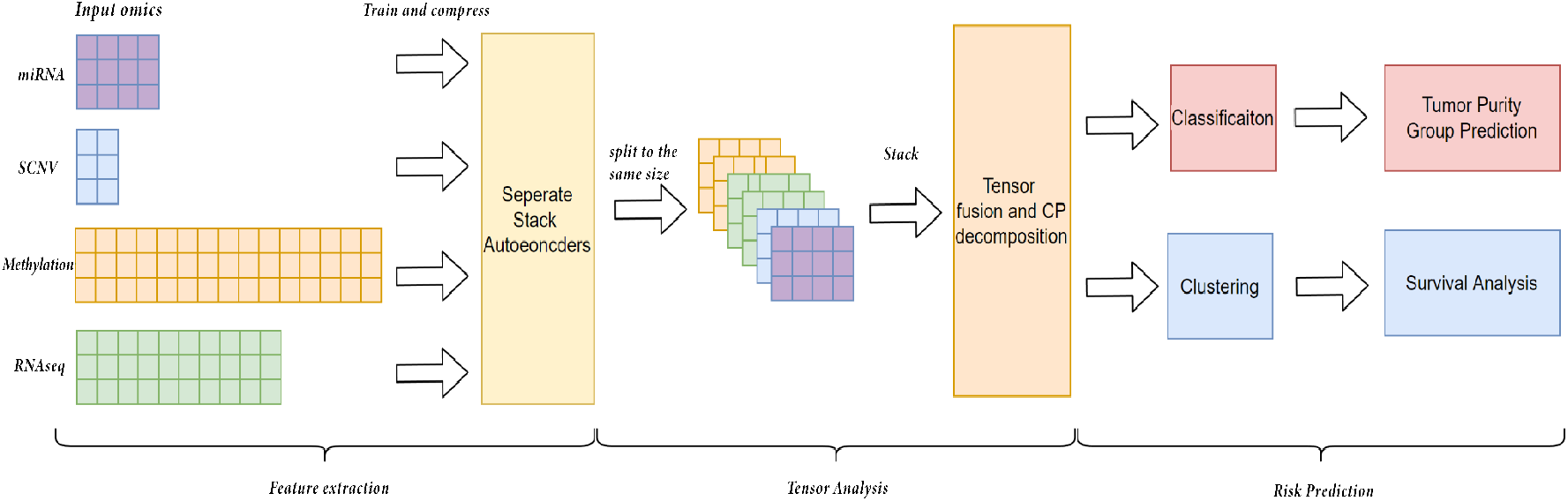
Our proposed framework contains three main components: feature extraction, tensor analysis, and risk prediction.

#### 3.2.1 Data Preprocessing

The collected datasets are split into 70% training data and 30% testing data to ensure a sufficient number of test cases. Data cleaning is applied to both breast and glioma datasets to handle the missing values. Only methylation contains a limited number of missing values, so we replace them with the mean value of the related gene features. Since it is uncertain whether zero values in omics data are meaningful or not, we decided to keep them to avoid any loss of meaningful information. After the cleaning phase, the data is scaled by the MinMaxScaler function as follows

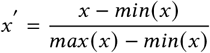

#### 3.2.2 Feature Extraction using Stacked Autoencoder

Since the sizes of omics data are various and can be extremely large due to many genes, it is necessary to reduce or compress them to a reasonable size. We aim to keep the maximum information in the extracted features from all the omics datasets. To achieve this goal, the stacked autoencoder model is implemented and applied to separate omics. It consists of an artificial neural network widely used for dimension reduction. It aims to extract meaningful information from the input dataset, transform them into smaller size latent and reconstruct the input data from the latent [Bank et al.(2020)]. To create the stacked autoencoder model, we have implemented the following steps:

##### Step 1: Encoding

Given an omics dataset, *D* with *N* samples and *d* features, an encoder in the autoencoder model compresses the *d* features into *d*′ where d >d’. The hidden layers stack within the encoder, reducing nodes between *d* and *d*′. The encoder part uses a non-linear mapping function to map the input data to hidden layer units and between the hidden layers. Assume *h* denotes the activation of the hidden layer neural unit, then its mathematical expression is as follows

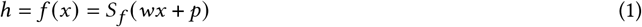

where *w* represents the learning weighted matrix connecting the input layer and the stacking hidden layers. *S*_*f*_ is the activation function at the last hidden layers, which is usually a Sigmoid function or a Tanh function as shown below in Eq. 2 and Eq. 3 respectively.

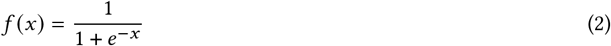

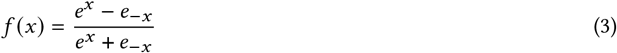

We also add a ReLU activation function in each hidden layer which has the following equation

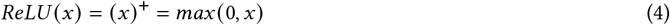

The following section shows the structure of the encoder for both datasets: Breast Invasive Carcinoma and Glioma.

##### Encoder Structure for Breast Invasive Carcinoma

After preprocessing, the four selected omics for breast data contain the same sets of samples and various features, e.g. SCNV, miRNA, RNAseq, and methylation, including 69, 823, 20155 and 335854 features, respectively. The structure will differ since they are all served by a separate autoencoder except for SCNV. The input features on the input layer are the initial features. Considering the size of SCNV is very small compared with others, we impute zero value to increase the size so that other omics do not need to be significantly reduced to match the specific dimension size. Therefore, SCNV was imputed to 512 features and fit into the autoencoder. Since the deep learning model may not learn zero value, the information will not be diluted [Mitra et al.(2020)]. The values for each layer are selected from a specific range so that the dimension of the current hidden layer is half of the last layer. Due to hardware limitations, it cannot set a higher value than 1024 on the first hidden layer for the two large omics. Hence, selecting the optimal target latent feature size below 1024 becomes necessary. More specifically, there is a ReLU activation function between each hidden layer. A three layers encoder is supplied with specific hidden nodes on each layer for the two large omics, while miRNA is served by a two layers encoder and SCNV is served by a single-layer encoder, as shown in Fig. 2

**Fig. 2.**
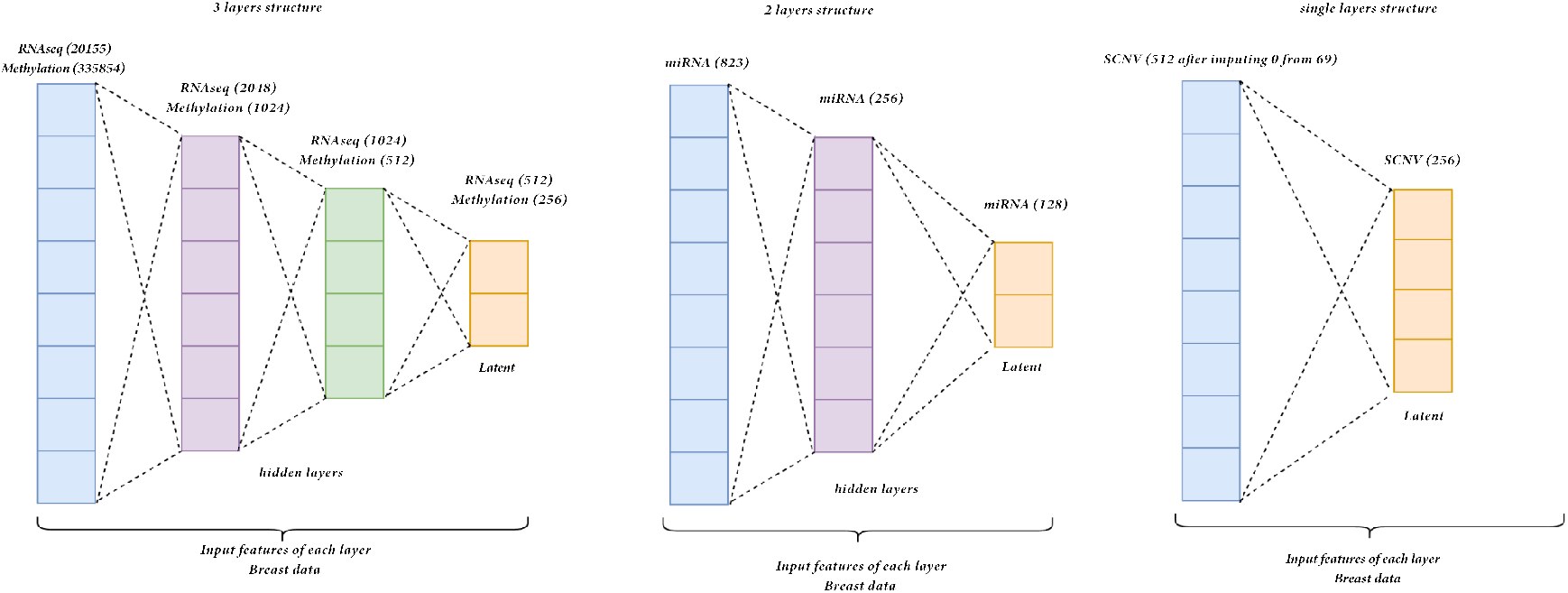
Encoder Structure for Breast Invasive Carcinoma

##### Encoder Structure for Glioma

The same four omics in the breast data are selected for glioma, with minor differences in the feature size. Initially, SCNV has 72 features, miRNA has 791, methylation has 336630, and RNAseq has 20118 features. Since these omics’ feature sizes are similar to those in breast data, encoders with similar structures are implemented for them. During the evaluation through training loss, there are changes in the target latent size and the features in the hidden layers for some omics. Same to the methylation in the breast, it cannot increase the output features to more than 1024 of the first hidden layers due to hardware limitations. Hence, it is compulsory to select the optimal target latent feature size below 1024. More specifically, there is a ReLU activation function between each hidden layer. Similar to the ones in breast data, the two large omics are served by the three layers encoder while the other two smaller omics fit into the two layers encoder. The detailed structure is presented in Figure 3.

**Fig. 3.**
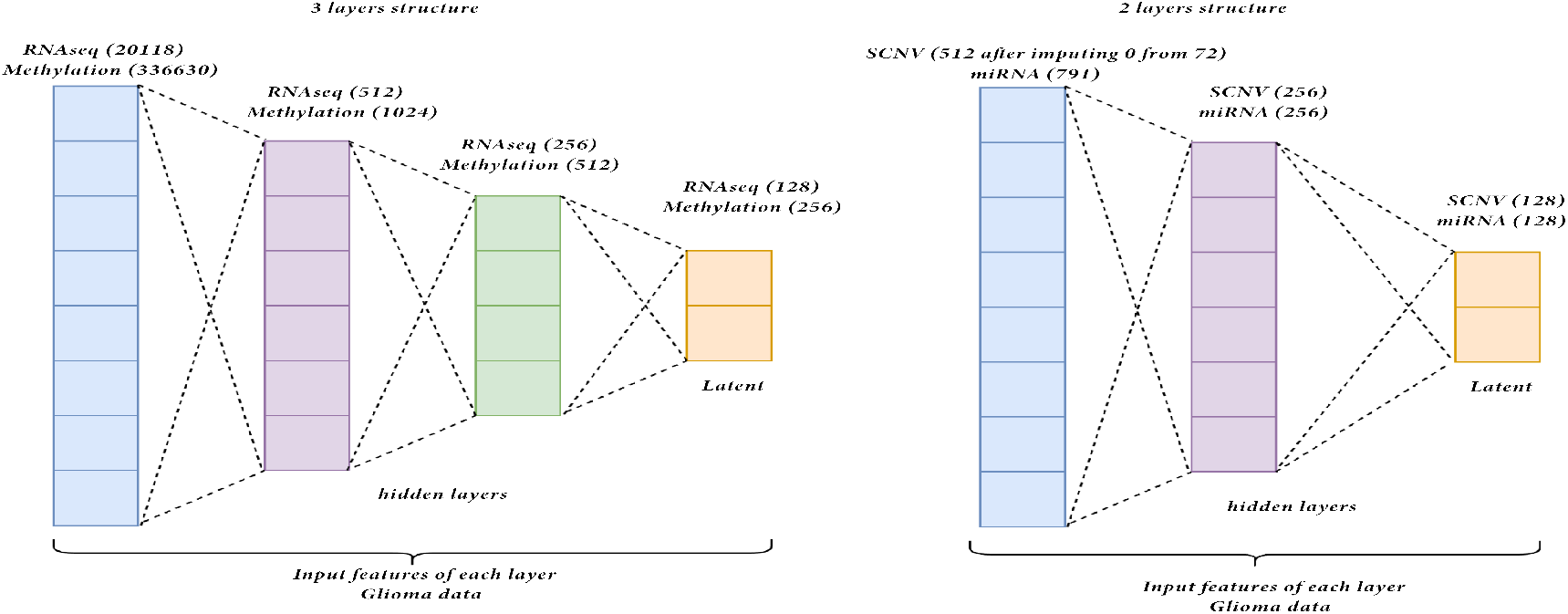
Encoder structure for Glioma

##### Step 2: Bottleneck

The compressed output is generated in the latent space in the bottleneck layer, having the same feature size as the number of nodes in the last hidden layer of the encoder. This latent output is regarded as the compressed output of the model. There are two usages of this latent output. The first usage is to put into the decoder of the stacked autoencoder to reconstruct the original input and evaluate the model by calculating the loss between the original input and reconstructed output. The other usage is to take this latent as the model output for the next component of our framework. For Breast Invasive Carcinoma, The optimal latent size for each is selected by inspecting the training loss and the validation loss in 10-fold cross-validation and gaining the one with the lowest training loss and stable low validation loss. After evaluating different targets, e.g. latent feature sizes, including 64, 128, 256, 512 and training the model using the entire train set, the resulting latent for each omics are 256 for SCNV, 128 for miRNA, 256 for methylation and 512 for RNAseq. For glioma, Similar to the breast cancer data, the evaluation of the optimal latent feature size is performed in a similar way using 10-fold cross-validation. As a result, the optimal latent for each omics is 128 for SCNV, 128 for miRNA, 256 for methylation and 128 for RNAseq.

##### Step 3: Decoding

The decoder part of the model mirrors the encoder part. Setting the same numbers of hidden layers, the decoder aims to reconstruct the input from the latent as follows

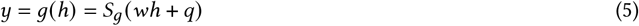

where the *w*′ represents the weight matrix between hidden layers, *y* represents the reconstructed input and *S*_*g*_ represents the activation function for the decoder.

##### Step 4: Loss function and back-propagation

To calculate the loss between the original input and reconstructed output, Mean Squared Error(MSE) is the loss function commonly used for autoencoder training. Assuming input *x* and target *y*, the loss can be written as

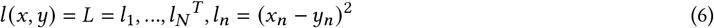

where *N* is the batch size 128. Since the default setup of the model is used

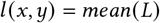

##### Parameter setting

The autoencoder is trained using 10-fold cross-validation to determine the optimal target latent size. Using the entire train set, each model will be trained again by setting the latent output as the optimal value. After running ten epochs, both the average training loss and validation loss of each model are around 1% to 3%. Adam is selected as the optimizer, and the learning rate is set to 0.001 to avoid overlearning. The latent for each test set is generated by the trained models.

#### 3.2.3 Multi-omics Tensor Data Fusion and Decomposition

##### Tensor Data Fusion

A 3D tensor is used to fuse these latents of each omics data. However, The matrices in the tensor must have the same size. To retain most of the information fused into the tensor, the latent embeddings with larger sizes are divided into multiple smaller embeddings with the same size as the smallest latent embedding. These matrices are the same size, so they can be stacked to form a tensor. The stacking strategy frequently merges multiple data sources into a single tensor that can be utilized in machine learning models. The effectiveness of this approach is impacted by the quality of the extracted features. If the features are noisy or not relevant to the intended task, then the stacking strategy may not be effective. However, in our method, we implement an autoencoder first to compress the data and learn new features using non-linear functions.

Four sets of latent features for breast training data are created, which contain the following shapes (samples, latent features) among various omics: SCNV (431,256), miRNA (431, 128), RNAseq (431, 512) and methylation (431, 256). The test data contains the same feature size with 185 samples. To integrate these latents into the same size, the minimum size among these latents is set as the target and split the larger ones evenly to the target. For example, we split four pieces of RNAseq having a shape (431,128) for each. Then, the pieces can be stacked to form a tensor, as demonstrated in Fig. 4. Therefore, it can fuse all the related data compressed by autoencoders into a tensor. After stacking them in the orthogonal axis, we successfully retrieve two tensors with shapes (431, 9, 128) and (185, 9, 128) for the train and test sets, respectively. Similar to the breast data, four latents belonging to glioma are generated after compressing the original by autoencoder. The shapes of each training set are as follows: SCNV (355,128), miRNA (355,128), RNAseq (355,128) and methylation (355, 256). The test sets share the same feature size separately and have 153 samples. After splitting to match 128, the minimum feature size, they are stacked in the orthogonal axis to form two tensors with shapes of samples, assays, and latent features, e.g., (355, 5, 128) and (153, 5, 128) for the train set and test sets respectively.

**Fig. 4.**
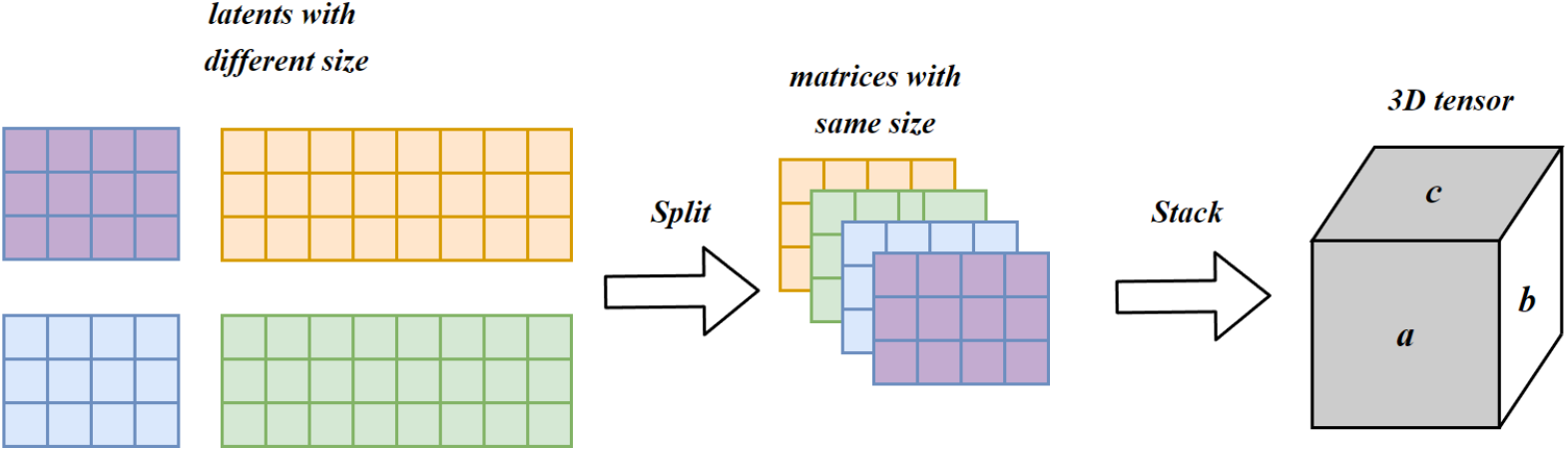
Stacking the matrices with the same size to build the tensor of shape: samples, assays, and latent features

##### Tensor Decomposition process

Given a tensor *X* ∈ *ℜ*^*I*×*J*×*K*^, We use Parafac [Carroll and Chang(1970)] (a.k.a CP decomposition) to decompose the tensor into three matrices A, B and C as shown in Fig. 5. Matrix A represents the patient’s mode, B represents the omics feature mode and C represents the genes (latent features) mode. In this sense, a tensor *X* can be written as

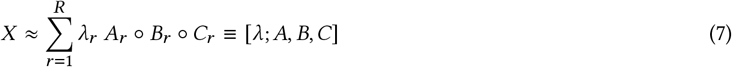

where “◦” is a vector outer product. *R* is the latent element, *A*_*r*_, *B*_*r*_ and *C*_*r*_ are r-th columns of component matrices *A* ∈ *ℜ*^*I*×*R*^, *B* ∈ *ℜ*^*J*×*R*^and *C* ∈ *ℜ*^*K* ×*R*^, and *λ* is the weight used to normalize the columns of *A, B*,and *C*.

**Fig. 5.**
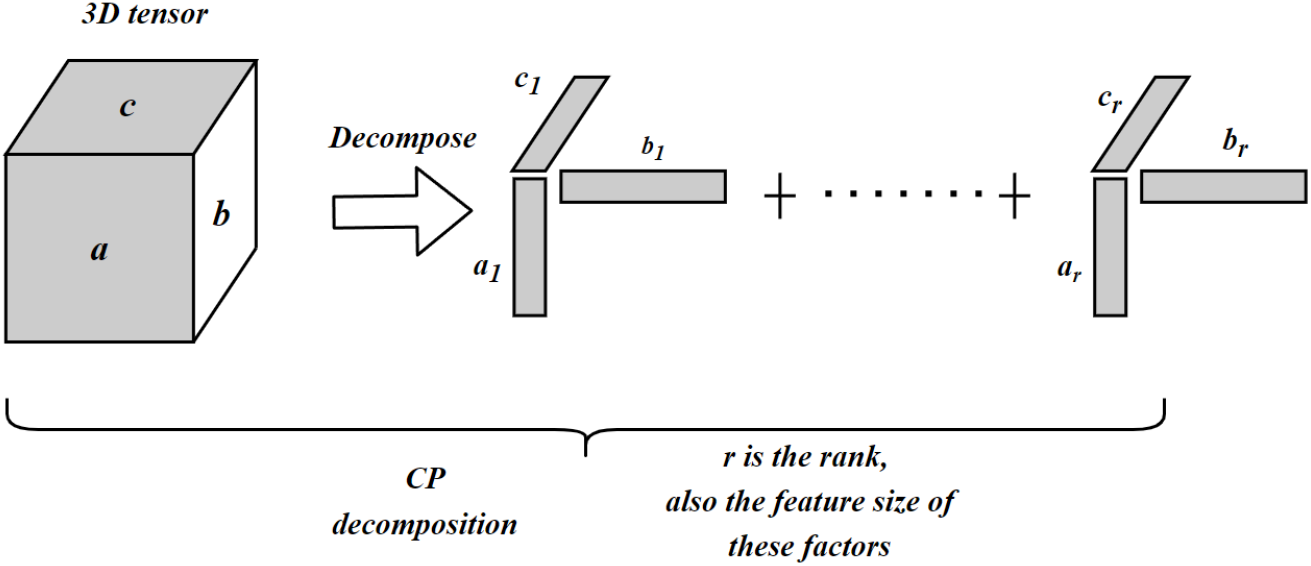
Tensor decomposition

The main goal of CP decomposition is to decrease the sum square error between the model and a given tensor *X* :

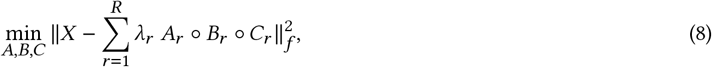

where 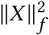 is the sum squares of *X*, and the subscript *f* is the Frobenius norm. In this work, we use the core consistency diagnostic technique (CORCONDIA) technique described in [Bro and Kiers(2003)] to determine the number of rank-one tensors *R* when it decomposed using the CP method.

It seems at first that the function presented in Equation 8 is a non-convex problem since it aims to optimize the sum squares of three matrices. However, the problem can be reduced to a linear least squares problem by fixing two of the factor matrices, and solving only the third one. Following this approach, the ALS technique can be employed which repeatedly solves each component matrix by locking all other components until it converges.

We remark that ALS can lead to sensitive solutions and it is not, in general, robust and hence motivates the need to incorporate the notion of penalty and regularization. Incorporating regularization and penalization parameters into the *L*_1_ norms makes it possible to achieve smooth representations of the outcome and thus bypass the perturbation surrounding the local minimum problem [Anaissi et al.(2018)]. The algorithm for CP decomposition using regularized ALS (RALS) is described in Algorithm 1. The *L*_1_ penalty terms 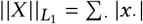 enforces the intensity of sparsity in *X*.

###### Algorithm 1: Regularized Least Squares for CP

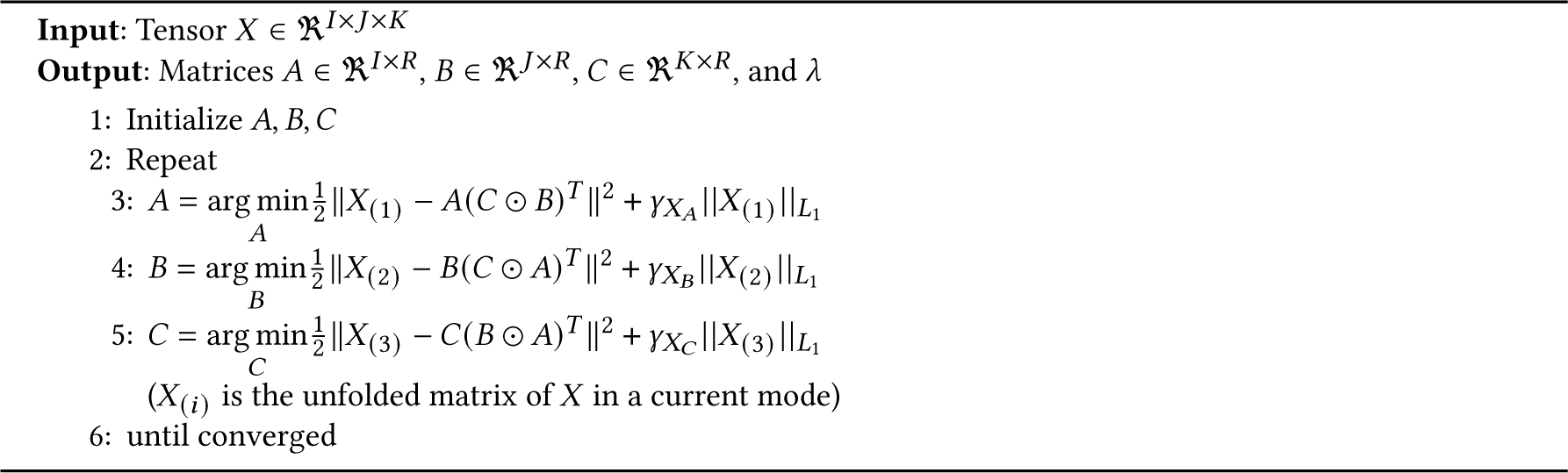

Interestingly, our proposed method employs a tensor for data fusion. The alternative naive approach would simply concatenate the multi-omics datasets into one single two-dimensional matrix. However, unfolding the data and analyzing them using two-way methods may lead to information loss since it breaks the modular structure inherent in the data. Therefore, a tensor data fusion approach will allow us to arrange the data from a set of multi-omics datasets as one single data structure T called a tensor. This tensor T has data in a form of a three-way tensor X ∈ R^*A*×*B*×*C*^ where *A* represents the number of multi-omics dataset, *B* represents the number of features in each omic dataset, and *C* is the total number of patients. The structure of this tensor is shown in Fig. 6

**Fig. 6.**
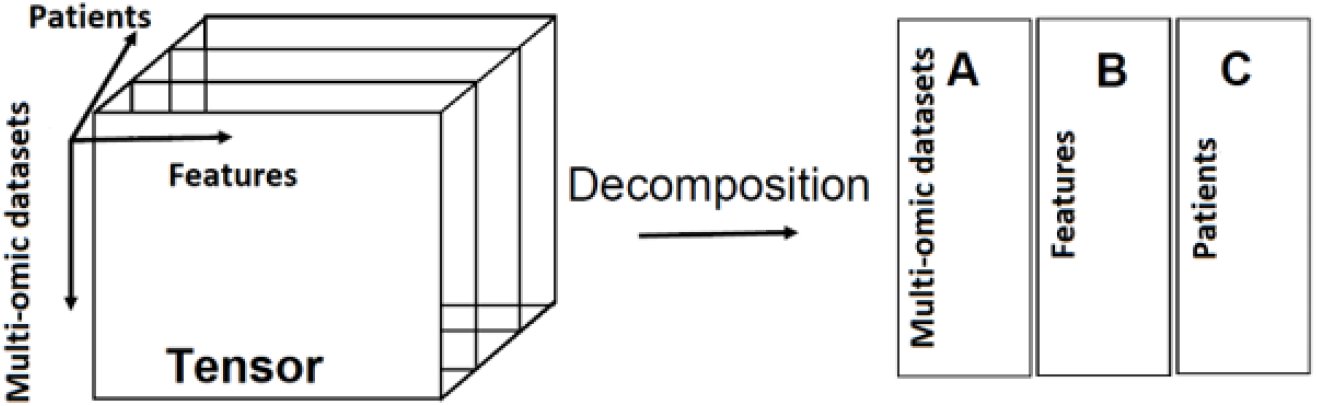
Multi-omics data fused in a tensor.

#### 3.2.4 Multi-omics Clustering and Prediction Models

In this section, the patients are stratified into low and high-risk groups using the latent features from the integrated multi-omics data: SCNV, miRNA, RNAseq, and methylation. To rationally identify different subsets of patients associated with different overall survival (OS), several clustering methods are investigated to cluster the patients into two groups using the integrated latent features from four omics including K-means, hierarchical clustering, DBSCAN, and Gaussian Mixture Model. To demonstrate a different performance of low and high-risk groups, prognostic significance is evaluated using univariate (Kaplan-Meier) and multivariate (Cox-regression) models across treated patients from the breast and glioma datasets. The p-value evaluates the statistically significant level. Further, tumor purity classification model is developed on the breast data as it is available only in the clinical breast data. Tumor purity is an important medical feature that explains the proportion of cancer cells. We categorize the tumor purity level into high and low levels based on the threshold of 0.7. The patient is considered as a high purity level when the tumor purity value is greater than or equal to 0.7 and low otherwise [Cheng et al.(2020)]. It is worth mentioning that the data is divided into the training and testing sets for the clustering and classification models.

## 4 EXPERIMENTS AND RESULTS

The evaluated datasets are downloaded from the public linkedomics repository^1^ including four single-omics datasets of SCNV, methylation, miRNA and RNAseq in addition to the clinical dataset for each cancer type. Breast Invasive and Glioma are the only two types with more than 600 clinical samples compared with all other cancer types. The chosen four single-omics datasets also have sample sizes of over 600, which are sufficient for the analysis. The core consistency diagnostic technique (CORCONDIA) suggests the rank *R* = 9 for the breast multi-omics tensor and 5 for glioma tensor [Bro and Kiers(2003)]. We compared our proposed method to MOFA [Argelaguet et al.(2018)].

### 4.1 Survival Analysis for Glioma

We investigate whether the patients can be stratified into risk groups for glioma cancer using the latent features from the multi-omics genomics data. The latent features are learned from our proposed framework, as shown in Fig. 1. First, for each type of cancer, the data is decomposed to 70% training data for model building and 30% testing data. Hierarchical clustering divides the patients into two or three risk groups. Then, to evaluate the ability of the multi-omics latent features to stratify patient overall survival (OS), a univariate regression model is fitted across glioma patients in the training set (N=330) and testing set (N=144). The significance levels are indicated as –*log*_10_ (p-value). Kaplan–Meier curves visualize the probability of survival outcomes over time in each group as shown in Fig. 7 and 8. A general observation is revealed from the results that multi-omics latent variables are significantly associated with patient OS in univariate models across all the patients in the training set. The patients could be stratified into low (N=147) and high-risk (N=183) groups and three groups with significantly different OS (p-value<0.05) as shown in Fig. 7.

**Fig. 7.**
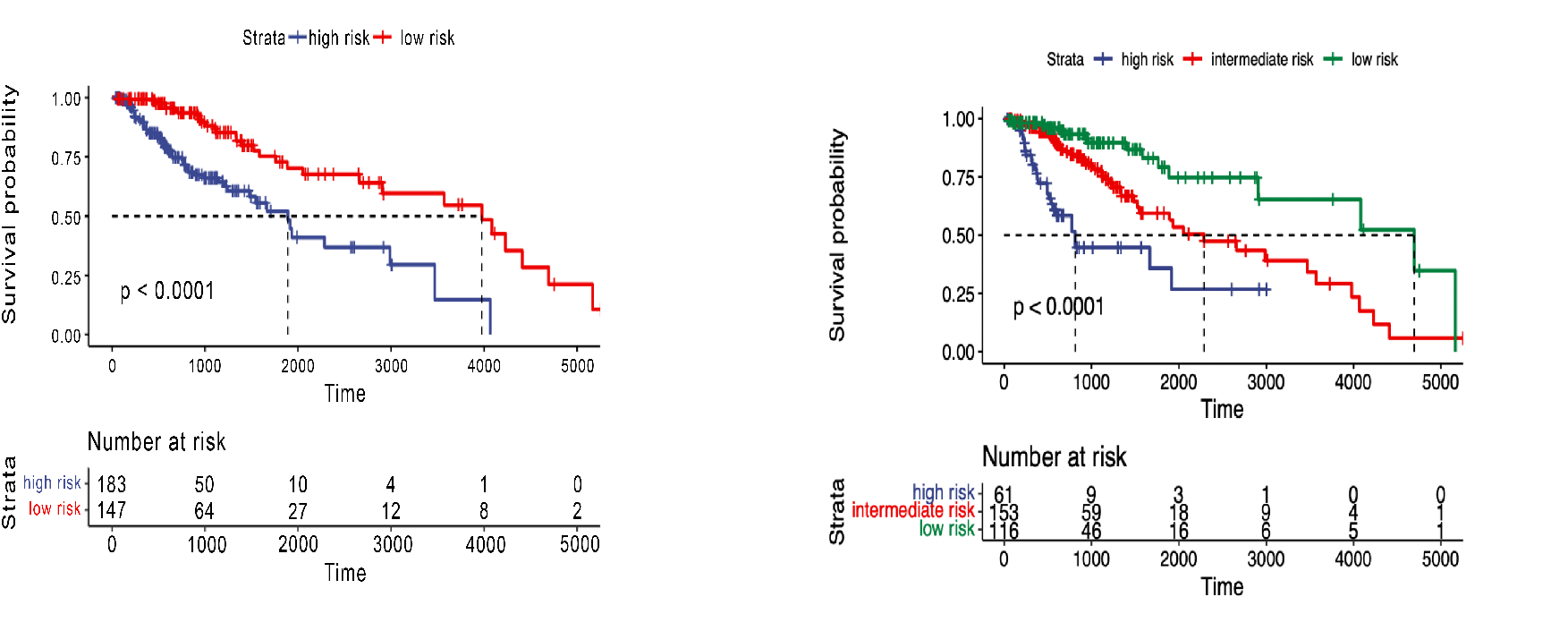
Overall survival of our method on glioma patients in training set stratified by hierarchical clustering using multi-omics latent variables. The ‘p’ value represents the P-value of the log-rank test comparing the different groups.

**Fig. 8.**
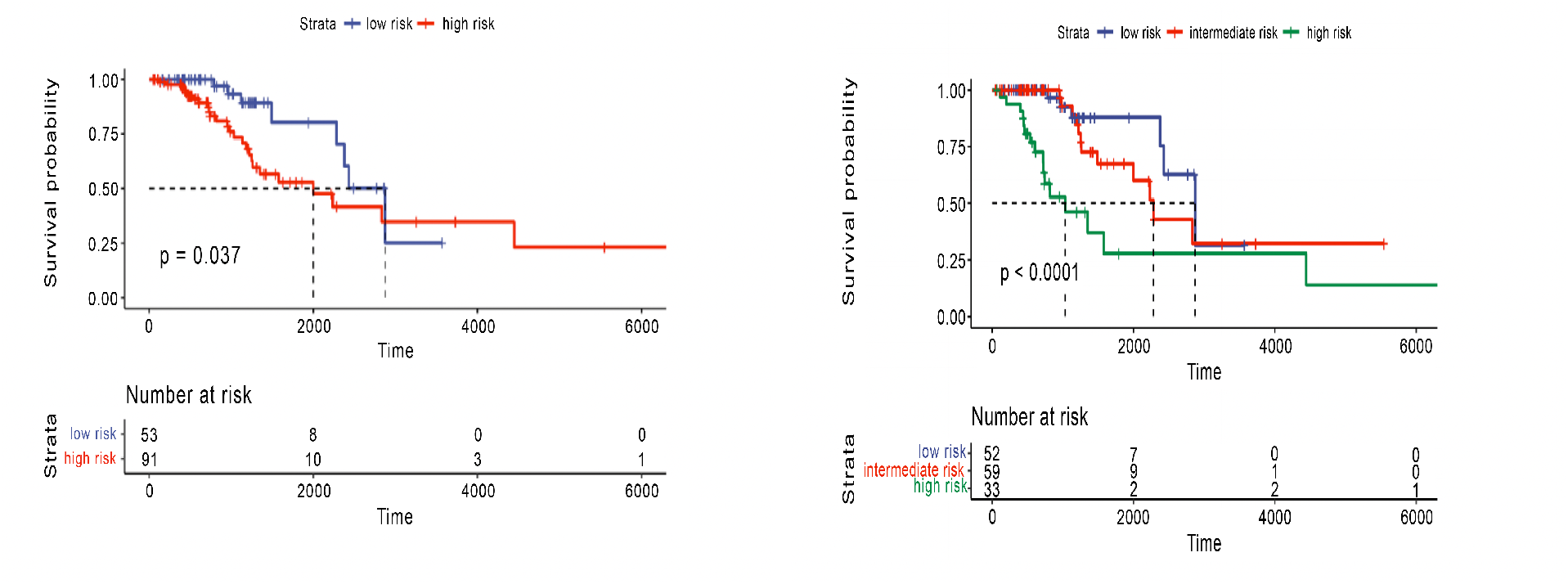
Overall survival of our method on glioma patients in the testing set stratified by hierarchical clustering using multi-omics latent variables. The ‘p’ value represents the P-value of the log-rank test comparing the different groups

For the test set of glioma cancer, both significant results are observed if cluster the data is into two groups and three groups (Fig.8). The p-value of 0.037 can be obtained when clustering the test set into two risk groups and less than 0.0001 for three risk groups. We compared our results with the state-of-the-art method, MOFA. The latent factors extracted by MOFA from both the training and testing sets did not significantly stratify patients into two or three risk groups, as demonstrated in Figs. 9 and 10. The p-value was not significant in all training and testing sets, except for the glioma testing set, which was significant. These results were obtained using five factors that resulted in the best performance using the MOFA method. Therefore, our framework can generate important latent features from multiple genomics data related to the patient’s overall survival. The clustering model can dichotomize patients with statistically significant p-value across all glioma patients.

**Fig. 9.**
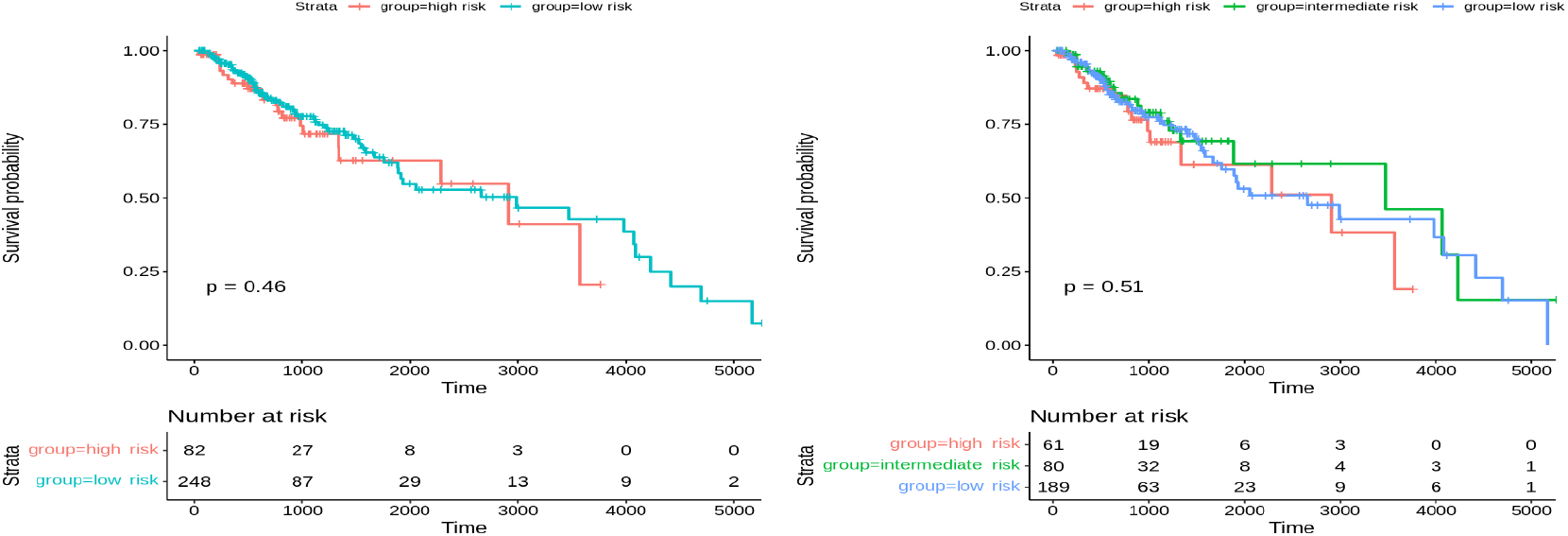
Overall survival of MOFA on glioma patients in training set using multi-omics latent variables. The ‘p’ value represents the P-value of the log-rank test comparing the different groups

**Fig. 10.**
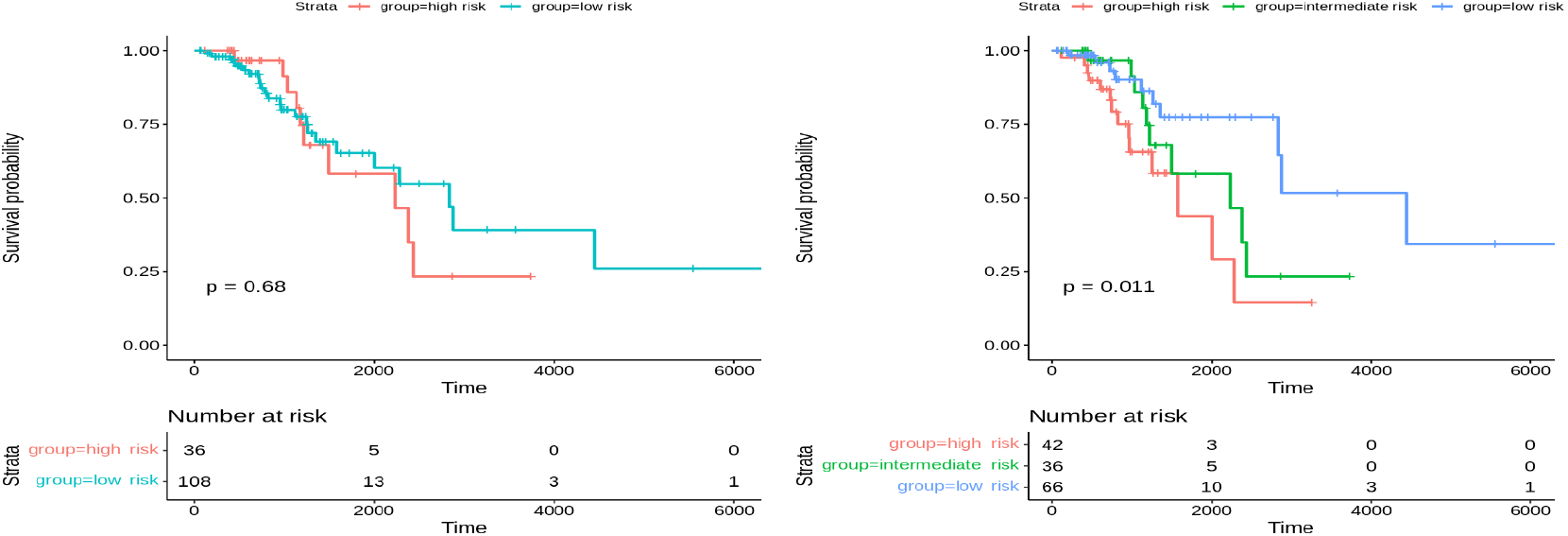
Overall survival of MOFA on glioma patients in the testing set using multi-omics latent variables. The ‘p’ value represents the P-value of the log-rank test comparing the different groups

### 4.2 Survival Analysis for Breast Cancer

The patients in the training set of breast cancer (N=426) using the learned latent variables from multi-omics data can be stratified into two groups and three groups by using the combination of Canberra distance and ward linkage for the hierarchical clustering algorithm with the significant difference between the two risk groups (p-value is 0.0085) and for three groups of p-value 0.029. The survival curves are shown in Fig. 11. However, the testing set (N=181) patients cannot be stratified into risk groups with significant differences using hierarchical clustering. The Kaplan-Meier survival curves of the risk groups do not show significant differences at the 5% significance level between the two and three curves (p-value is 0.078 and 0.16, respectively), as shown in Fig. 12. Our method outperformed MOFA in significantly stratifying breast cancer patients into multiple risk groups. This was observed by developing a clustering model that dichotomized breast cancer patients using the latent factors of the MOFA method. As shown in Figs. 13 and 14, there was no statistically significant difference between the two and three risk groups across all breast cancer patients in both the training and testing sets.

**Fig. 11.**
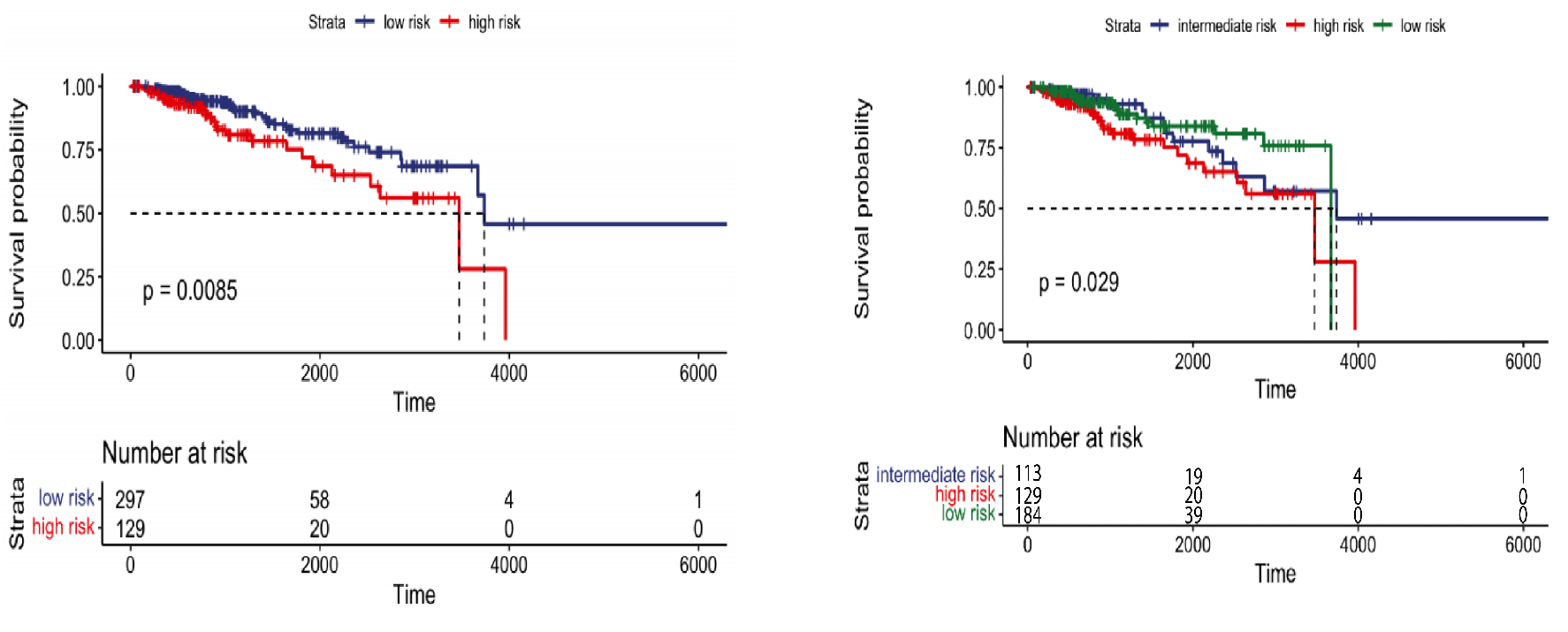
Overall survival of our method on breast patients in training set stratified by hierarchical clustering using multi-omics latent variables. The ‘p’ value represents the P-value of the log-rank test comparing the different groups.

**Fig. 12.**
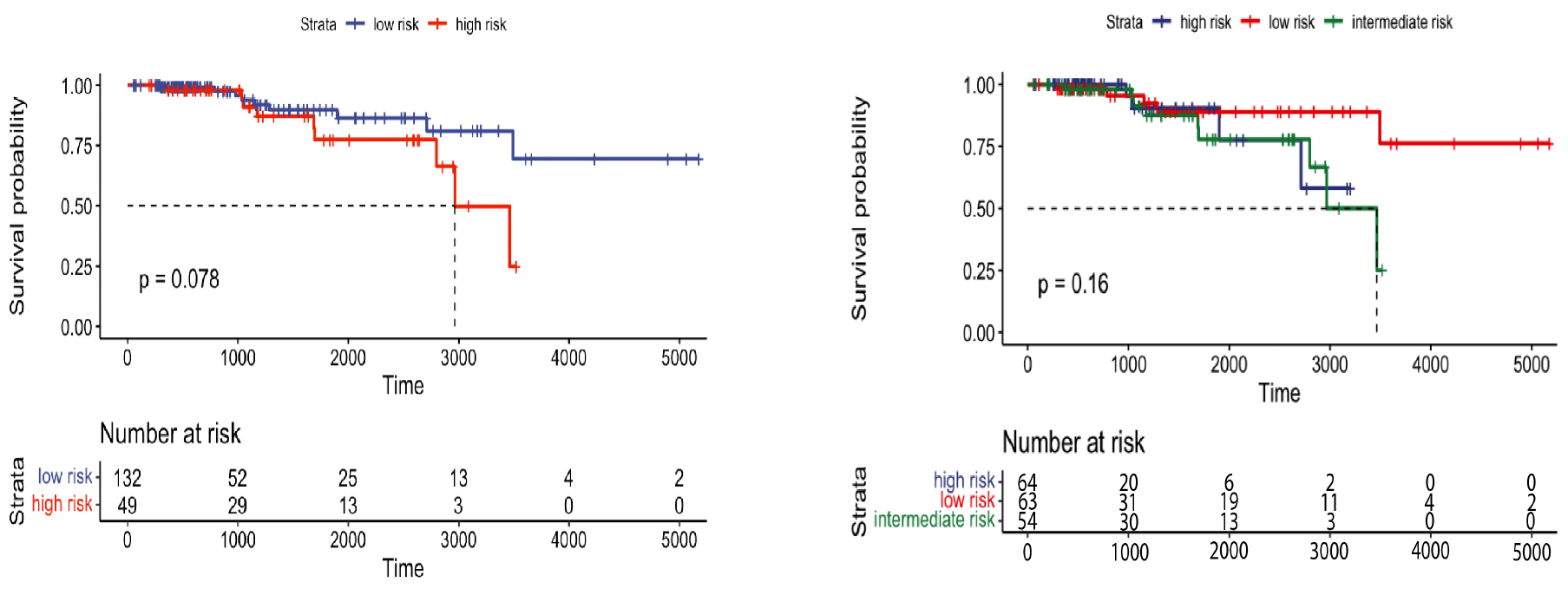
Overall survival of our method on breast patients in testing set stratified by hierarchical clustering using multi-omics latent variables. The ‘p’ value represents the P-value of the log-rank test comparing the different groups

**Fig. 13.**
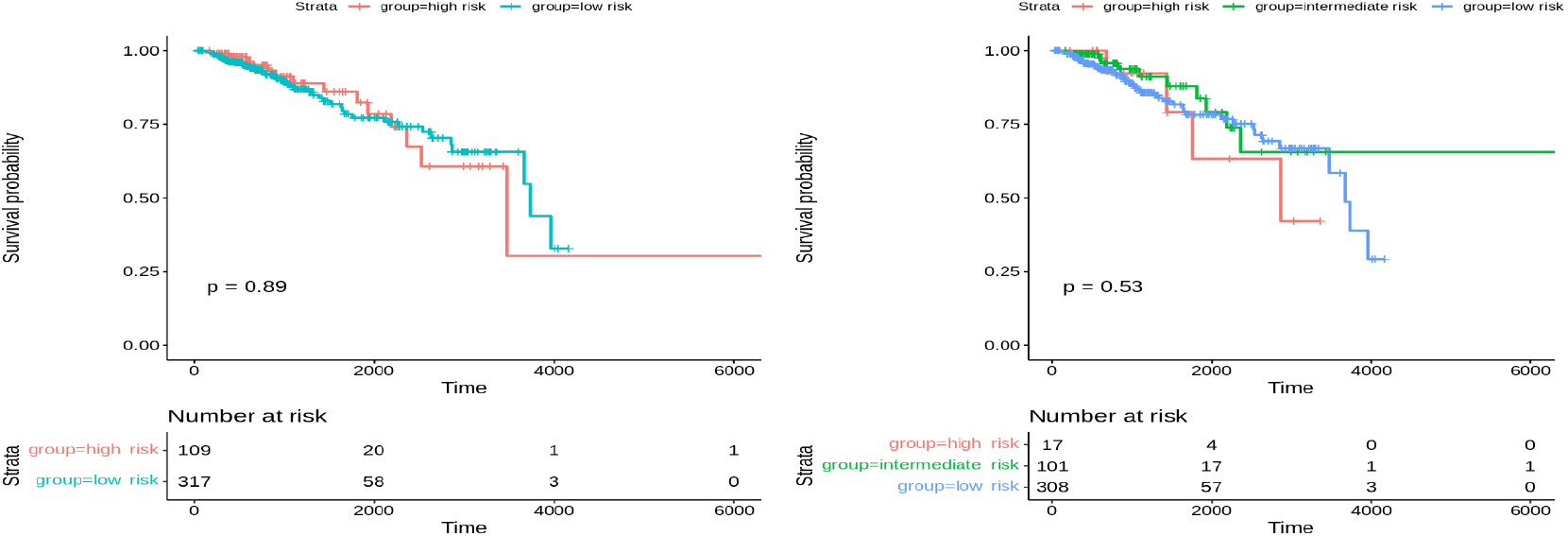
Overall survival of MOFA on breast patients in training set stratified by hierarchical clustering using multi-omics latent variables. The ‘p’ value represents the P-value of the log-rank test comparing the different groups

**Fig. 14.**
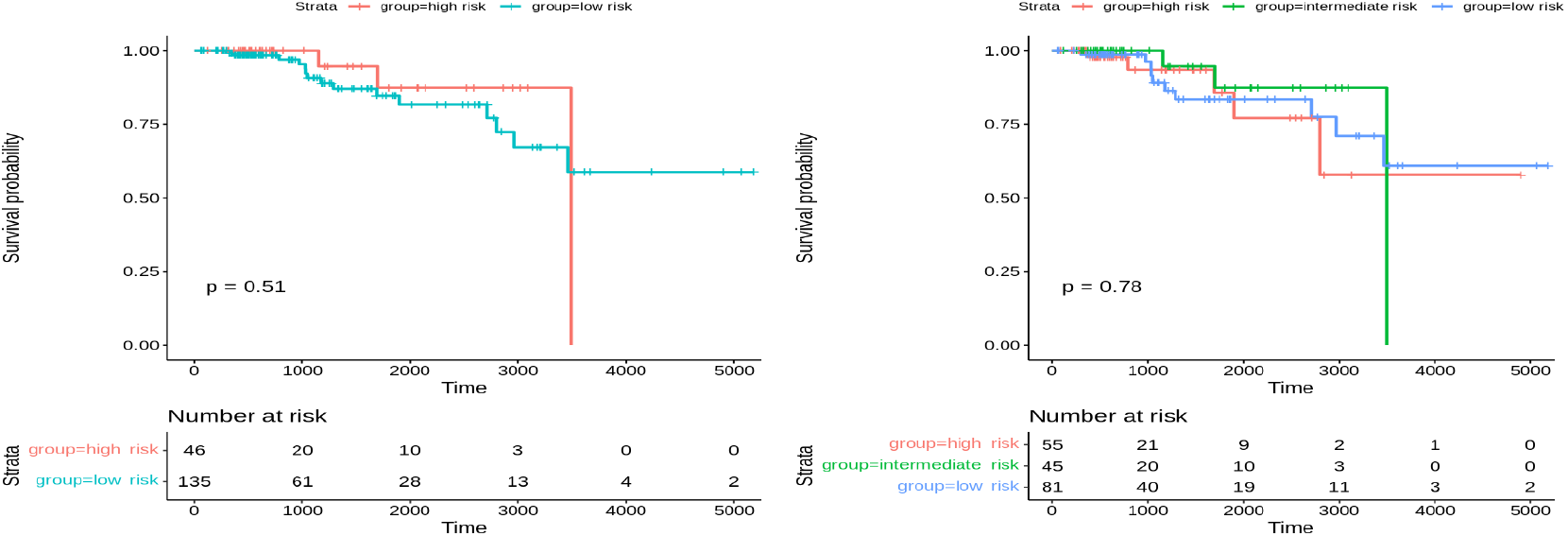
Overall survival of MOFA on breast patients in testing set stratified by hierarchical clustering using multi-omics latent variables. The ‘p’ value represents the P-value of the log-rank test comparing the different groups

Since very few patients can survive longer than 3000 days, to achieve more significant results, we restrict the survival time of patients to up to 3000. When adopting the combination of maximum distance and ward linkage for the hierarchical clustering algorithm, both training and test sets’ results are significant. As shown in Fig. 15, the p-value of 0.015 is observed when clustering the train set of breast cancer into two groups, while the p-value of 0.032 is obtained when clustering the test set of breast cancer into two groups.

**Fig. 15.**
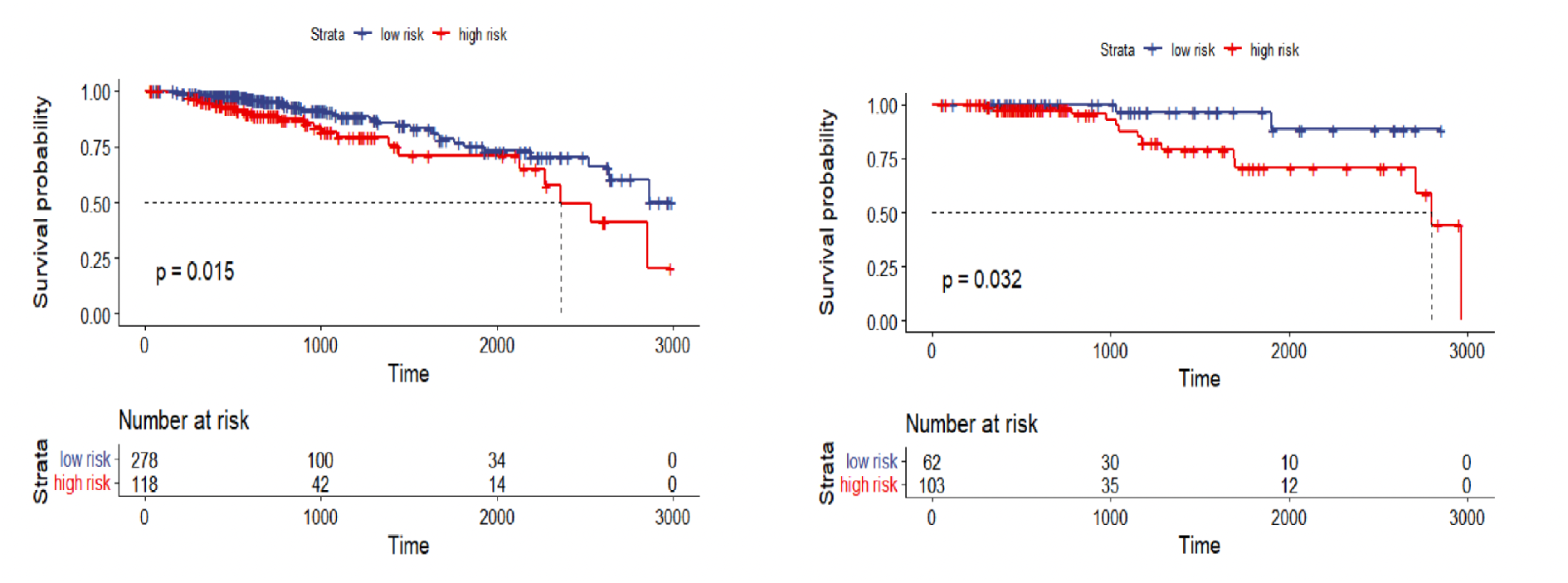
Overall survival of our method on breast patients in the training and testing sets with restriction survival time to 3000 days which stratified by hierarchical clustering using multi-omics latent variables. The ‘p’ value represents the P-value of the log-rank test comparing the different groups.

Overall, the results for glioma cancer perform better than the ones for breast cancer. The results demonstrate that the features extracted from the autoencoder models are significant after tensor decomposition, which further proves the utilization of the multi-omics data is meaningful for determining the risk of patients with a specific cancer type. By identifying patients’ risk levels, the overall survival rate is expected to be increased by either involving earlier inventions for cancers or choosing better therapies for patients with different stages of tumor.

### 4.3 Interpret Latent Variables using t-SNE Visualization

Deep learning methods have shown remarkable success in our method to learn the non-linear representation of the data. However, one of the main challenges with deep learning methods is the interpretation of the learned features, including latent variables, which can be highly complex and abstract, making it difficult to interpret the meaning of individual latent variables. Latent variables represent underlying biological features that cannot be directly observed, but visualization techniques such as t-SNE can be used to interpret them graphically.

In this experiment, we first applied t-SNE on the latent variables extracted from our proposed method to identify clusters of samples with similar latent variable values. As shown in Fig. 16, the resulting clusters can provide insights into the underlying biological processes or molecular pathways driving differences between samples. For example, they may suggest that the latent variables from multi-omics are capturing differences in gene expression or other molecular features associated with the disease subtype. Next, we evaluated the latent variables generated by MOFA for multi-omics. The results, visually observed in Fig. 17, demonstrate that our proposed method more effectively separates samples using multi-omics latent variables in the glioma testing dataset.

**Fig. 16.**
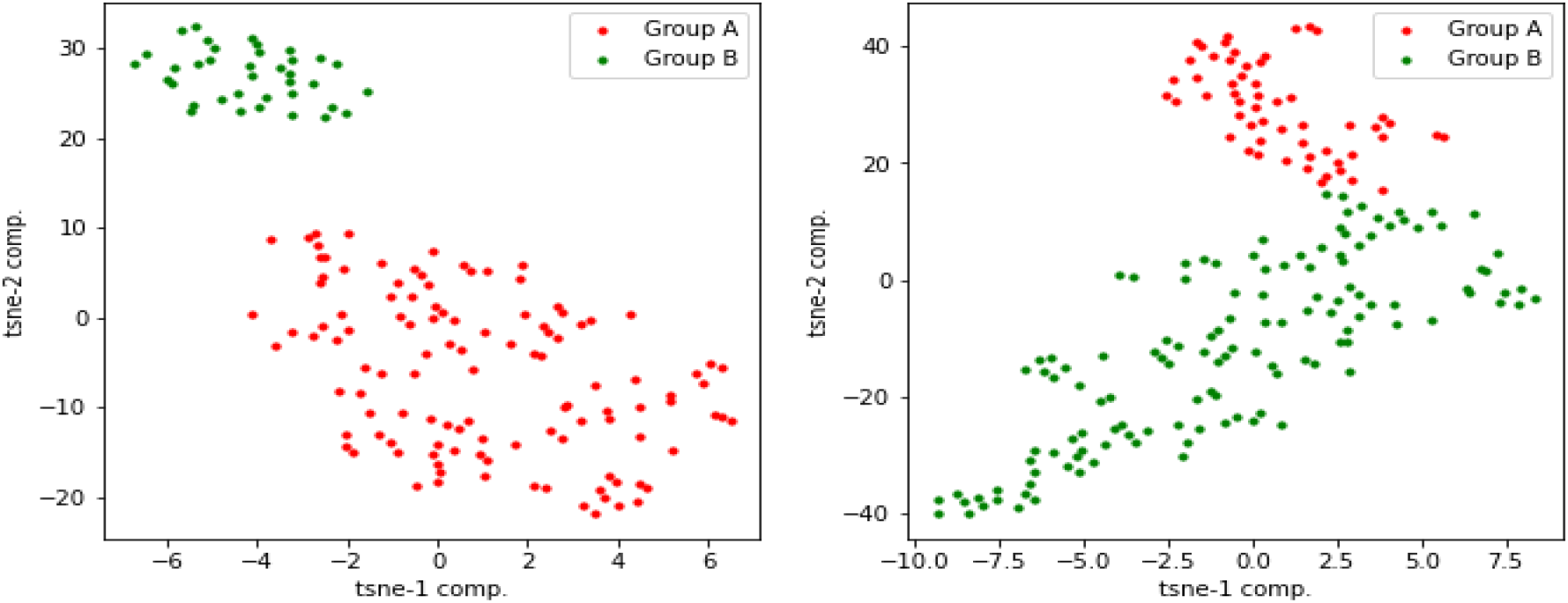
t-SNE plots of multi-omics latent variables in our method for glioma and breast testing data respectively.

**Fig. 17.**
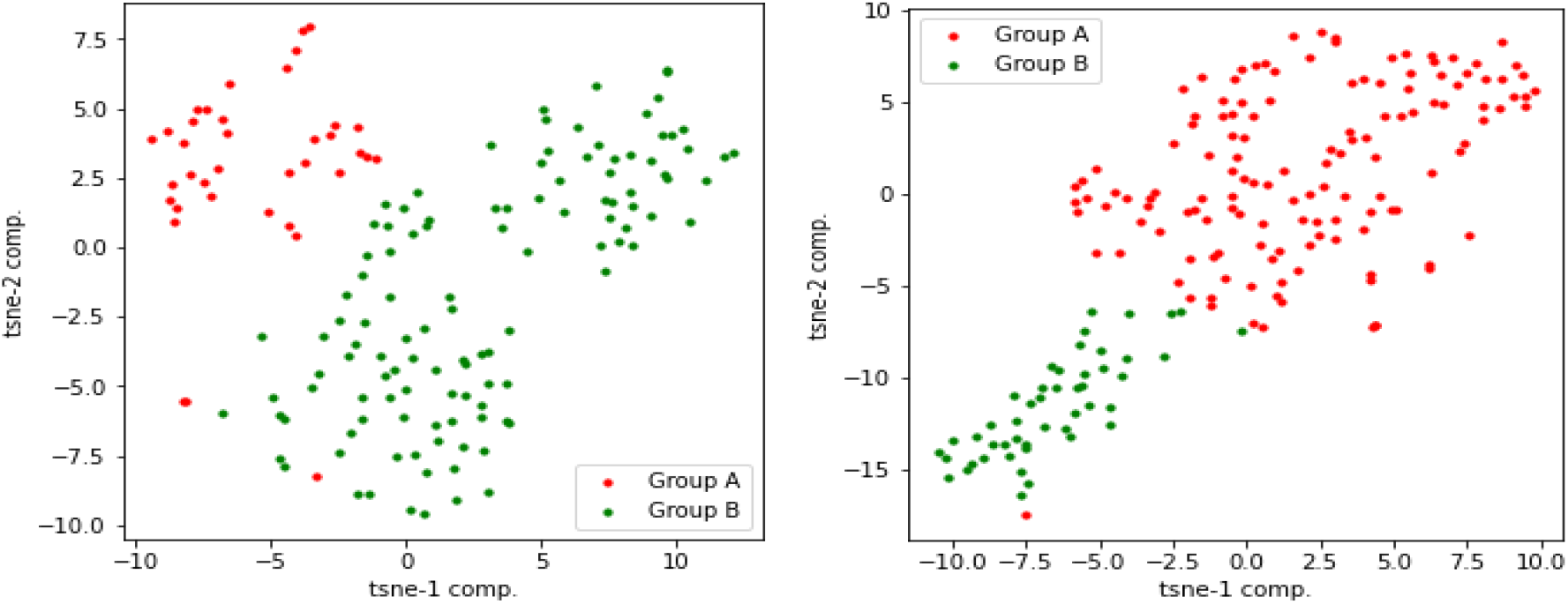
t-SNE plots of multi-omics latent variables in MOFA for glioma and breast testing data respectively.

**Fig. 18.**
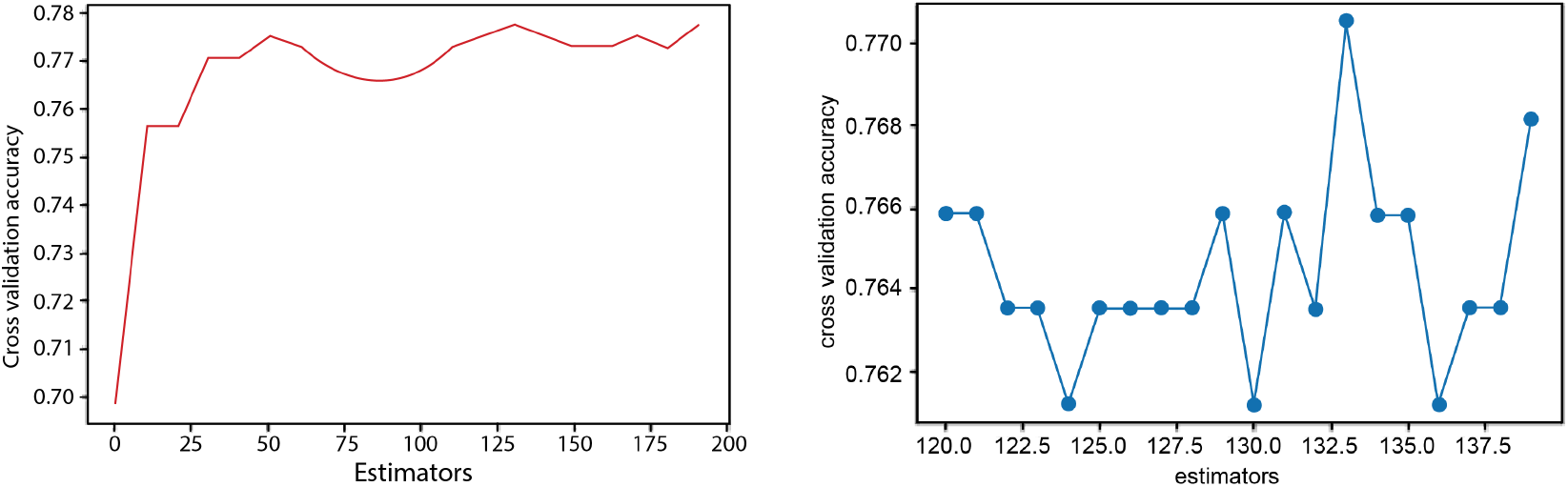
Parameter tuning.

### 4.4 Classification on Tumor Purity for CP decomposition of Breast

We further evaluate the multi-omics latent variables to classify the patients based on tumor purity as presented in Table 1. We have conducted the experiments on the breast data only because this feature exists only in its clinical data. Tumor purity is categorised into low and high groups based on a threshold of 0.7. Comparing the performance of different classification models based on the result sets of CP decomposition and tucker decomposition, it is observed that the model trained with the data after CP decomposition performs better. This classification problem is particularly challenging in the breast dataset, and even state-of-the-art methods struggle to achieve high accuracy [Li et al.(2019)]. Specifically, by testing the result sets of CP decomposition with different ranks, the best model is Random Forest which is trained on the result sets of CP decomposed by setting the decomposed rank to 9. The accuracy rate is 0.69, and the weighted F1-score is 0.65. For the remaining models, Logistic Regression, Naive Bayes, SVM, and AdaBoost have the same accuracy of 0.59, but their F1-score does not reach 0.4, so their performance is relatively poorer. Therefore, Random Forest can be used to classify tumor purity if using the CP decomposition method.

**Table 1.** Classification of Tumor Purity on CP Decomposition.

|  | Accuracy | Macro F1 | Weighted F1 |
| --- | --- | --- | --- |
| **Logistic regression** | 0.59 | 0.37 | 0.44 |
| **KNN** | 0.41 | 0.29 | 0.23 |
| **Naive Bayes** | 0.59 | 0.37 | 0.44 |
| **Decision tree** | 0.41 | 0.29 | 0.23 |
| **SVM** | 0.59 | 0.37 | 0.44 |
| **Random forest** | **0.69** | **0.60** | **0.65** |
| **Gradient Boosting** | 0.44 | 0.36 | 0.32 |
| **Adaboost** | 0.59 | 0.37 | 0.44 |

Three parameters are tuned in the random forest model: estimators, max depth, and max features. Among them, estimators have the most significant impact on the results. The optimal values are identified using the search space. Firstly, we search estimator values from 0 to 200. A random forest is built based on the interval of 10, and the intervals are taken as the x-axis, and the corresponding cross-validation scores are set as the y-axis (Fig.18). Based on the results, the highest accuracy value when the estimator is 133. Finally, using the estimators of 133 as the determining parameter, optimal values of both max depth and max features can be obtained in a similar process, which is two and nine, respectively.

## 5 DISCUSSION

We first indicate the significance of using multi-omics data and fusing the latent variables using tensors to identify the cancer risk groups. In this section, we conduct experiments using single-omics data on breast cancer. The patients are clustered into two groups, and the survival analysis is conducted separately to see whether those features extracted from autoencoder models of single-omics are significant or not. The p-value is reported in Table 2, which shows the insignificance difference between risk groups by using the single-omics separately for all technologies for breast data. On the other hand, the survival analysis results on glioma single-omics data are significant as presented in Table 3. Therefore, the performance of the latent variables of single-omics datasets of glioma is consistent with the latent variables of multi-omics, which proves that using the features from multiple omics produces significant results in both breast and glioma datasets.

**Table 2.** P-value to stratify the patients into two risk groups for training and testing breast data on single-omics.

|  | Training | Testing |
| --- | --- | --- |
| **RNAseq** | 0.25 | 0.28 |
| **SCNV** | 0.58 | 0.43 |
| **miRNA** | 0.15 | 0.58 |
| **Methylation** | 0.48 | 0.094 |

**Table 3.** P-value to stratify the patients into two risk groups for training and testing glioma data on single-omics.

|  | Training | Testing |
| --- | --- | --- |
| **RNAseq** | <0.001 | 0.013 |
| **SCNV** | <0.001 | 0.004 |
| **miRNA** | <0.001 | 0.04 |
| **Methylation** | <0.001 | 0.005 |

Overall, this study delivers a novel framework that overcomes few potential issues. Firstly, the framework handles every single-omics separately instead of combining all the omics at the first step. Concatenation-based integration methods, which combine all the omics into a single large matrix, have proved to integrate the multi-omics information in some cases (Reel et al. 2021). However, one of the main issues is that the information in smaller omics is very likely to be lost. In our case, SCNV is at high risk of missing information because it only has 69-72 feature size, while methylation is far larger than it, with more than 330k features. The framework approached in this study can avoid this concern as it handles each omic separately before fusing. Implementing autoencoders individually compresses as much as the original information of each single-omics. Secondly, instead of forcing a common target dimension for all the omics to reduce to, compressing each of them to their most optimal size makes more sense since the aim is to keep the maximum information of single-omics during the compressing process. Again, combining multi-omics to a large matrix can lose the information from the smaller size omics. It may be missing information when the feature extraction or selection methods are implemented on this large integrated matrix. Integrating these multi-omics after they were compressed by a separate autoencoder can reduce the risk of information loss, as these multi-omics data will no longer have huge size differences. The framework compresses the maximum information for each omic and then integrates them into a tensor, minimizes the information loss brought by compressing as a whole, and avoids handling multi-omics on integration when they have huge size differences.

Objectively, the framework does not add any knowledge from the biological area. We aim to investigate the biological interpretation of the difference between the low and high-risk groups identified by the latent variables extracted from multi-omics cancer data which could include:

- Differential activation of oncogenic pathways: The latent variables may be capturing differences in the activation of pathways involved in cancer development and progression. Patients in the high-risk group may have higher levels of activation of oncogenic pathways, leading to more aggressive tumor growth and a worse prognosis.
- Immune system dysfunction: The latent variables may be associated with differences in the immune response to cancer. Patients in the high-risk group may have immune system dysfunction, such as reduced immune surveillance or an immunosuppressive tumor microenvironment, which allows the tumor to evade detection and destruction by the immune system.
- Treatment response: The latent variables may be predictive of how well patients will respond to different cancer treatments. Patients in the high-risk group may be less responsive to standard treatments, leading to a worse prognosis.

## 6 CONCLUSION

We propose a multi-omics framework using deep-learning autoencoders and tensors to identify the cancer risk groups. Multi-omics integrates diverse omics data including methylation, somatic copy-number variation (SCNV), micro RNA (miRNA) and RNA sequencing (RNAseq). Our proposed framework use autoencoders for each omics data separately to reduce the number of dimensions and capture the maximum information. The latent variables are extracted from individual omics data and integrated using tensors, then, the common features are identified using CANDECOMP/PARAFAC (CP) decomposition. The low-dimensional multi-omics data is clustered into two and three risk groups using Hierarchical clustering. Several survival analysis experiments have been conducted which indicated that the low dimensional multi-omics data can be stratified into high and low-risk groups. Further, a classification model is constructed using the fused features from multi-omics data to predict the tumor purity in breast cancer. The future direction of this work will involve incorporating biological knowledge to further investigate the inter-relationships among different techniques and molecules.

## Data Availability

Publicly available
https://linkedomics.org/login.php

## Footnotes

1 http://linkedomics.org

